# Machine learning for estimating and comparing clinical rules for treating diarrheal illness with antibiotics

**DOI:** 10.1101/2025.01.10.25320357

**Authors:** Allison Codi, Sara Kim, Elizabeth Rogawski McQuade, David Benkeser, the AntiBiotics for Children with severe Diarrhea (ABCD) Study Group

## Abstract

Acute diarrheal disease is one of the leading causes of death in children under age 5, disproportionately impacting children in low-resource settings. Many of these cases are caused by bacteria and therefore could respond to antibiotic treatment; however, the benefits of widely prescribing antibiotics must be weighed against the risks for the emergence of microbial resistance. These challenges present the opportunity for developing individualized treatment guidelines for diarrheal disease. In this study, we utilize a framework for the creation and evaluation of individualized treatment rules that leverage diagnostic and other clinical information to recommend antibiotic treatment to children with watery diarrhea. In contrast to many applications of pipelines for creating and evaluating treatment rules, we (i) explicitly consider creating rules that limit the proportion of children treated under a rule, to limit risks for overtreatment and the emergence of microbial resistance and (ii) propose methods to compare the performance of rules based on different sets of input covariates, which allows for quantification of the impact of measuring additional diagnostic biomarkers in clinical settings. We use a nested cross validation procedure that makes use of ensemble machine learning and doubly-robust estimation approach to derive, evaluate, and compare rules. We demonstrate that our proposed method yields appropriate inference in a realistic simulation study and apply our method to a real-data analysis of the AntiBiotics for Children with severe Diarrhea (ABCD) trial.

## 1 INTRODUCTION

Interest in personalized medicine has grown in recent years due to the realization that treatment guidelines may need to be tailored to specific individuals in order to maximize the individual- and/or population-level benefit while minimizing risks. One specific research area where developing individualized treatment guidelines is critical is in the management of acute diarrheal illness with antibiotics. Acute diarrheal disease accounts for approximately 450,000 deaths in children under age 5 each year, with a disproportionate burden borne by children in sub-Saharan Africa and South Asia ^1^. Diarrheal episodes can lead to complications including increased risk of future mortality ^2^ and growth faltering ^3,4^. Antibiotics are effective in treating diarrhea caused by bacterial pathogens; however, antibiotics are not expected to be efficacious against diarrhea caused by viruses or protozoa. Moreover, antibiotic use is a primary driver of antibiotic resistance, making it essential to limit inappropriate use, particularly for viral diarrhea episodes ^5^. However, because rapid pathogen diagnostics are unavailable at the point of care in many settings, treatment guidelines are based on insensitive syndromic criteria which leads to both antibiotic overuse and missed opportunities to appropriately treat episodes that could benefit. The value of point-of-care diagnostics to guide treatment decisions relative to the value of other clinical and epidemiological characteristics is unknown, but has important implications for the prioritization of diagnostic development. These challenges highlight the need to develop and evaluate rules for administering antibiotics to children with diarrhea using available data sources. Specifically, this evaluation must (i) consider different sets of input characteristics so that the relative benefits of adding rapid diagnostics can be weighed against potential costs and (ii) strike a balance between potential clinical benefits and the risk of antibiotic resistance.

Myriad statistical approaches have been proposed for developing individualized treatment rules based on a set of input characteristics. Many of these rules are defined by using standard machine learning algorithms to estimate the conditional average treatment effect (CATE), the average causal effect of treatment on the outcome of interest within subgroups defined by the selected set of input characteristics for the rule. These methods include the S, T, X, R, and doubly robust (DR) learners ^6,7,8^. Tree-based methods have also been developed for CATE estimation ^9,10,11^, among other approaches. Once one of these methods has been used to estimate the CATE, a treatment rule can be defined by thresholding the estimated CATE at a particular level – that is, the rule treats all individuals with estimated benefit beyond a chosen threshold, and does not treat any individual with expected benefit less than the chosen threshold.

Many existing applications of these methods are not fit for the purpose of developing rules for treating diarrheal illness in that standard applications generally consider (i) developing rules for a single, comprehensive set of input covariates, rather than comparing rules developed based on different sets of input covariates and (ii) thresholding the CATE at zero, thereby recommending treatment to all individuals who are not expected to be harmed by the treatment. In this study, we build on existing frameworks to tailor the approaches to the setting of developing rules for antibiotic treatment of diarrheal illness. Specifically, we constrain our rules to assign treatment only to individuals with a clinically relevant effect size and explore the trade-offs in terms of expected benefits for treated children compared to the risks of overuse of antibiotics. We also explicitly compare the performance of rules based on different sets of input covariates used to define the rule, thereby providing a way to quantify relative benefits of adding additional diagnostic tools to clinical settings. Comparing rules allows interrogation of the most informative covariates for treatment decisions, which can then be prioritized for data collection and incorporation into treatment decision-making.

Towards these goals, we utilize a cross-validation-based framework for developing and evaluating our treatment rules. Specifically, using repeated training samples, we use the DR learner to estimate the CATE ^12^, ensuring accurate CATE estimation if at least some combination of nuisance parameters are consistently estimated. We utilize ensemble machine learning, or super learning to develop estimates of nuisance parameters and the CATE ^13^. The super learner builds an asymptotically optimal ensemble of candidate models and has been shown to uncover treatment effect heterogeneity more effectively than traditional approaches ^14,15^. After CATE estimates are obtained, a candidate rule is defined by thresholding the CATE estimates, as described above. Using validation samples, we estimate various quantities that describe the performance of the rule. We specifically consider estimating the proportion of the population treated under the rule, as well as treatment recommendation-specific and population-level effects under the rule. Our estimators of these effects also leverage doubly robust approaches, ensuring robustness to model misspecification. We utilize the calculus of influence functions to propose methods that compare the performance of different rules in order to identify which sets of input covariates are most informative for treatment decisions. We demonstrate our method is approximately unbiased and has appropriate confidence interval coverage through simulations that closely mimic data from a randomized control trial (RCT) of an antibiotic for acute watery diarrhea in children. Lastly, we apply the method to data from this RCT to quantify important tradeoffs between strategies to target treatment of children with diarrhea. The method implementation used to run all analyses can be found in the *drotr* R package on GitHub ^16^.

## 2 BACKGROUND, NOTATION, AND PARAMETERS OF INTEREST

### 2.1 Motivating example: AntiBiotics for Children with severe Diarrhea (ABCD)

AntiBiotics for Children with severe Diarrhea (ABCD) was a clinical trial that examined the effect of azithromycin, a recommended antibiotic for diarrhea, on mortality and linear growth among children with non-bloody diarrhea ^17^. The trial enrolled children ages 2-23 months who presented with acute watery diarrhea and were undernourished or dehydrated across seven sites in Bangladesh, India, Kenya, Malawi, Mali, Pakistan, and Tanzania. Information collected upon enrollment included clinical characteristics, malnutrition indicators, and sociodemographic characteristics (refer to table 1 for full list of covariates). The first approximately 1000 children enrolled at each site had fecal samples collected that underwent qPCR pathogen testing to determine etiology of their diarrhea. This resulted in a total of 6,692 children with valid qPCR results to be included in the analysis. After enrollment, children were randomized 1:1 to a 3-day course of azithromycin or placebo. The primary outcomes of interest were all-cause mortality up to ninety days after enrollment and linear growth faltering ninety days after enrollment. Diarrheal status day three after enrollment was measured as a secondary outcome. There were 269 (4.0%) children missing the linear growth faltering outcome and 1857 (27.7%) children missing the day three diarrhea outcome. While the initial study did not find an overall survival benefit, there was a small improvement in linear growth that could not be explained by bacterial etiology alone, suggesting that a broader range of factors than etiology may determine treatment benefit ^2^. Incorporating additional factors such as clinical, malnutrition, and sociodemographic characteristics into treatment guidance may better target treatment and maximize benefit. This provides the motivation for our analysis, wherein we consider the impact of treatment rules that consider different sets of input variables.

**TABLE 1.**
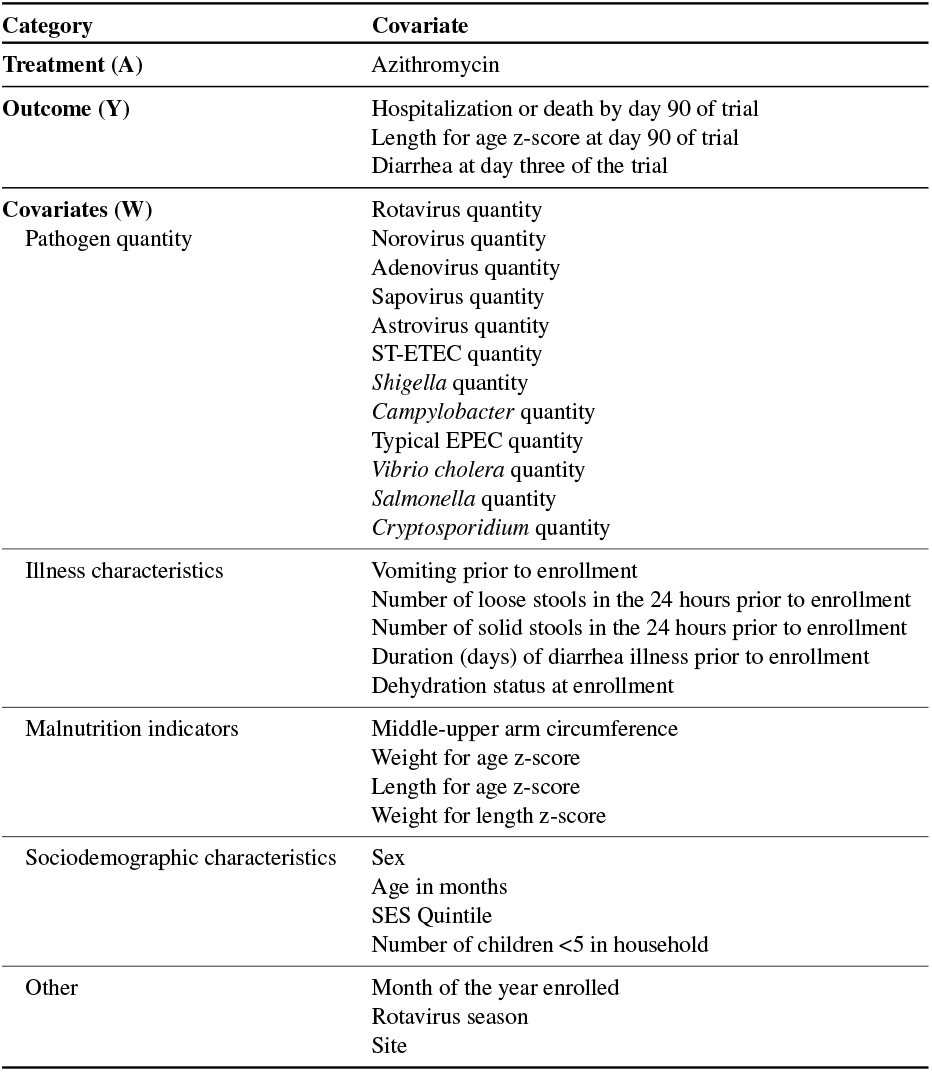
List of covariates and abbreviations

### 2.2 Notation

Let *W ∈ 𝒲* denote a set of measured baseline covariates. Let *Z ⊆ W* be a subset of covariates we are interested in using to create a treatment rule. Let *A ∈* {0, 1} indicate a binary treatment decision, which is equal to 1 if the participant is assigned azithromycin or equal to 0 if the participant is assigned placebo. Let *Y ∈ 𝒴* be a real-valued outcome variable of interest. In the ABCD trial, the outcomes of interest include hospitalization or death by day ninety of the trial (binary), diarrhea at day three of the trial (binary), and length-for-age z-score at day ninety of the trial (continuous). We simply use *Y* to denote any one of these outcome measures. Let Δ *∈* {0, 1} represent a missingness indicator for the outcome variable *Y*, which is equal to 0 if the outcome is missing and equal to 1 if the outcome is observed. We observe an independent and identically distributed sample of observations *O*_*i*_ = (*W*_*i*_, *A*_*i*_, Δ_*i*_, Δ_*i*_*Y*_*i*_) *∼ P*_0_ *∈ 𝒫*, where *P*_0_ is an unknown distribution in model *𝒫* that is non-parametric up to assumptions defined in Section 2.4.

For each treatment assignment *a ∈* {0, 1}, we introduce the counterfactual outcome *Y*(*a*, 1), which denotes the outcome that would have been observed if, contrary to actual assignments, a given individual were assigned to treatment *a* and there was no missingness in the outcome.

### 2.3 Treatment rules

Given *Z ⊆ W*, the CATE is defined as *ψ*(*z*) = *E*[*Y*(1, 1) | *Z* = *z*]–*E*[*Y*(0, 1) | *Z* = *z*] and describes the expected benefit associated with azithromycin treatment relative to placebo in a subpopulation defined by covariates *Z*. For example, if we are considering the day ninety hospitalization or death outcome and *Z* consists only of a single binary variable indicating the presence of blood in stool, then *ψ*(1) represents the benefit of azithromycin in children with blood in their stool, while *ψ*(0) represents the benefit for children without blood in their stool.

The CATE can be used to define a treatment rule *d* : *𝒵 →* {0, 1}, where it is common to consider rules of the form *d*(*Z*) = *I*(*ψ*(*Z*) *≤* 0), where *I*(*·*) is the indicator function. Such rules assign treatment only to all subgroups of individuals who are not harmed by the treatment on average. Such rules are not immediately relevant in our setting because it is unlikely that azithromycin treatment would be harmful for any children with respect to hospitalization or death and thus the optimal treatment rule would likely treat all children irrespective of the magnitude of benefit from treatment. We instead consider treatment rules that only recommend treatment to individuals that have expected treatment benefit of at least magnitude *t*, e.g., rules of the form *d*(*Z*) = *I*(*ψ*(*Z*) *≤ t*) for some *t* < 0. The goal of this approach is to identify subgroups that experience the greatest clinical benefit under the treatment rule and target this subgroup for treatment, thereby balancing the benefits of treatment with concerns of antibiotic overuse.

Note that the rule *d* depends on two features: (i) the selected set of covariates *Z* and (ii) the selected threshold *t*. For simplicity, we do not include any explicit notation to indicate the reliance of the rule on (i) and (ii); however, it is important to note this dependence as we describe approaches for comparing different rules in Section 2.4.

#### Remark

An alternative approach to controlling the amount of antibiotics prescribed under a rule would be to develop a so-called “resource-constrained” treatment rule ^18^. This approach is typically used when treatment is not widely available and it is of scientific interest to deliver the limited amount of treatment that is available to those who would benefit most from it. Though in the present setting azithromycin is generally widely available, this approach could be considered as a means of controlling the proportion of children who are prescribed azithromycin. Under this approach, an *a-priori* threshold could be selected for the proportion of children with watery diarrhea who would be prescribed antibiotics. For example, we may wish to develop a rule that prescribes antibiotics to no more than 50% of cases. The resource constrained rule would then recommend treatment to children with *ψ*(*Z*) below its median value, i.e., the 50% of children with the largest treatment effects. While this approach may be interesting to consider, treatment policies for diarrheal antibiotic use are unlikely to be made with resource constraints in mind. We argue that our approach is more patient-centered, in that we allow treatment to be given to anyone who is likely to have significant benefit from the treatment.

### 2.4 Effect Estimands for Evaluating Treatment Rules

Let *Y*(*d*) denote the counterfactual outcomes that would be observed if everyone in the trial had been treated according to treatment rule *d*, i.e., *Y*_*i*_(*d*) = *d*(*Z*_*i*_)*Y*_*i*_(1, 1)+{1–*d*(*Z*_*i*_)}*Y*_*i*_(0, 1). To quantify the impact of a particular rule *d*, we may be interested in the effect of the treatment on the outcome in the subgroup that is recommended treatment under rule *d, E*[*Y*(1, 1) – *Y*(0, 1) | *d*(*Z*) = 1]. We term this quantity the *Average Treatment effect among those Recommended Treatment by rule d* (*ATRT*_*d*_). Similarly, we may be interested in the effect of treatment on the outcome in the subgroup that is *not* recommended treatment under rule *d, E*[*Y*(1) – *Y*(0) | *d*(*Z*) = 0]. We term this quantity the *Average Treatment effect among those Not Recommended Treatment by rule d* (*ATNRT*_*d*_). By comparing these two quantities (*ATRT*_*d*_ – *ATRNT*_*d*_), we can assess the effectiveness of the treatment rule in distinguishing between children based on their expected benefit.

We are also interested in the proportion of individuals who are recommended treatment under a particular rule, *E*{*d*(*Z*) = 1}, termed the *proportion treated*, which allows us to evaluate treatment rules based on the expected number of children that would be recommended antibiotics by the rule. Weighing the proportion treated against the expected benefit in those treated allows for an explicit evaluation of the trade-off between maximizing clinical benefit and controlling the over-prescription of antibiotics.

Given the *ATRT*_*d*_ and the proportion treated, we can also quantify the population-level effect of rule *d, E*[*Y*(*d*) – *Y*(0, 1)], which compares the average outcomes at a population level if all children were treated according to rule *d* vs. no one was treated. We term this quantity the *Average Treatment effect under Rule d* (*ATR*_*d*_). We note that *E*[*Y*(*d*) – *Y*(0)] = *E*[*Y*(*d*) – *Y*(0) | *d*(*Z*) = 1] *× P*(*d*(*Z*) = 1) + *E*[*Y*(*d*) – *Y*(0) | *d*(*Z*) = 0] *× P*(*d*(*Z*) = 0) = *E*[*Y*(*d*) – *Y*(0) | *d*(*Z*) = 1] *× P*(*d*(*Z*) = 1), since *E*[*Y*(*d*) – *Y*(0) | *d*(*Z*) = 0] = 0, necessarily.

We are also interested in comparing effects under treatment rules based on different sets of input characteristics. For example, for two rules *d*_1_ and *d*_2_ developed based on the same fixed threshold, but different sets of input covariates *Z*_1_ *⊆ W* and *Z*_2_ *⊆ W*, we may wish to estimate 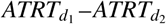. For example, we may set *Z*_1_ to be a limited set of covariates that are readily available in a clinical setting (e.g., age, sex, other demographic variables), while *Z*_2_ might include these readily available covariates but additionally include a set of diagnostic variables. The estimand 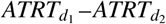 then provides a quantification of the additional benefit of measuring diagnostics for tailoring treatment guidelines. Comparing different treatment rules in this way allows us to identify whether and to what extent additional covariate measurements could help better tailor treatment of children. Identifying the most relevant subset of information needed to adequately inform treatment decisions would allow public health officials to allocate resources effectively while maximizing patient benefit.

For a given rule *d*, identification of *ATRT*_*d*_, *ATNRT*_*d*_, and *ATR*_*d*_ requires the following assumptions.

(A1) *Conditional randomization/conditional exchangeability:* There exists a set of covariates *W* such that *Y*(*d*) *⊥ A* | *W*.
(A2) *Stable Unit Treatment Values Assumption (SUTVA):* (A2.1) consistency such that for *a* = 0, 1, *E*[*Y*(*a*, 0) | *W*] = *E*[*Y*(*a*, 0) | *A* = *a, W*] = *E*[*Y* | *A* = *a, W*] and (A2.2) no interference such that *Y*(*a*_*i*_) *⊥⊥ a*_*j*_ for *i ≠ j*.
(A3) *Positivity:* (A3.1) treatment positivity under rule *d* such that *P*_0_{0 < *P*_0_(*d*(*Z*) = 1 | *W*) < 1} = 1 and (A3.2) missingness positivity such that *P*_0_{0 < *P*_0_(Δ = 0 | *A, W*)} = 1.
(A4) *Missingness at random:* The set of selected covariates *W* is such that *Y ⊥⊥* Δ | *A, W*.

Assumptions (A1) and (A4) imply that there is a sufficiently rich set of covariates *W* such that we can de-confound both the treatment and missingness probabilities. (A1) should be satisfied by design in the ABCD trial given that treatment was randomly assigned. (A4) is more difficult to ensure by design, as participant drop out is not always under control of the investigator. However, the majority of missingness of length for age z-score outcomes in the ABCD study can be attributed to the fact that some study sites began enrolling participants prior to to obtaining IRB approval to collect these data. After IRB approval was obtained, these data were collected on almost all participants (96%). Thus, we expect it is reasonable to assume that this outcome should be missing at random conditional on study site.

Assumption (A2.1) is sometimes referred to as the assumption of there being only a single form of treatment. Given that the treatment in question is a well-defined pharmaceutical intervention, we expect that this assumption is plausible in our context. Assumption (A2.2) asserts that there is no causal interference between individuals in the study. While this assumption is often dubious in the context of infectious disease studies, we expect that it is plausible in the context of ABCD given the large catchment areas for enrollment at each site and exclusion criteria for enrollment which included having a sibling currently enrolled in ABCD. Moreover, we expect that because the trial population is defined by children who present with diarrheal illness, the relevant impact of interference would be limited to additional infections acquired after enrollment into the trial, which is possible, but unlikely.

Assumption (A3.1) of positivity implies there is a non-zero chance of being treated under rule *d* for all sets of covariates *W*. This assumption is again satisfied by design for ABCD. Assumption (A3.2) stipulates that conditional on the selected set of covariates *W*, there is a positive probability of measuring the outcome of interest. Given the low levels of missingness, we again expect this assumption to be reasonable in the context of ABCD.

Under (A1)-(A4), we can identify our effects of interest for a summarizing a given treatment rule *d* as follows:

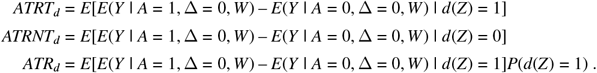

### 2.5 Data-adaptive effect parameters

The goal of our analysis is to simultaneously develop and evaluate treatment rules for different sets of input variables *Z*. Because the same data set will be used both for treatment rule development and for evaluating the rules, we rely on cross-validation to deliver appropriate inference. Specifically, we utilize *K*-fold cross-validation wherein a given input data set is partitioned randomly into into *K* splits of approximately equal size that are used to define *K folds* where each fold consists of a *training sample* and a *validation sample*. In the *k*-th fold the validation sample is defined as the *k*-th partition of the data, while the training sample is defined as the remaining *K* – 1 partitions.

Our procedure uses the *K* training samples to derive *K* distinct treatment rules. The corresponding validation samples are then utilized to estimate the effects of interest. Finally, estimates of effects are averaged over the *K* folds to arrive at an estimate of the average performance of the *K* rules.

By averaging over cross-validation folds, our procedure is targeting a data-adaptive target parameter ^19^. To formalize our target of inference, we introduce the notation 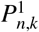 to denote the empirical probability distribution of the *k*-th validation sample and 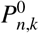 to denote the empirical distribution of the *k*-th training sample. The *k*-th training sample is used to create treatment rule 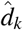 (for details see Section 4.3 below). We then consider this rule as fixed and given, and go on to estimate effects based on this rule in the validation sample. For (*a, d*) *∈* {0, 1}^2^, let 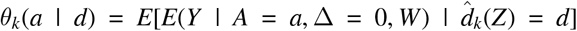 and let 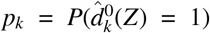. Thus, under our assumptions 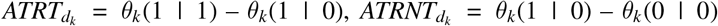 and 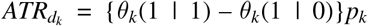. We can formally define our target of inference by averaging over cross-validation folds. Thus, for example, we consider inference on the average *ATRT* across the *K* rules learned in the *K* training samples,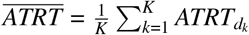.

Our focus on data-adaptive effect parameters is in line with previous work in this area, and previous results have established that in large samples, inference based on an asymptotic Normal distribution approximation should lead to appropriate conclusions ^19^. It is also interesting to compare our data-adaptive parameters to effect estimates under optimal treatment rule for a given threshold *t* given by *d*^*\**^(*Z*) = *I*(*E*[*Y*(1, 0) – *Y*(0, 0) | *Z*] > *t*). We expect that as sample size increases, if our training sample specific rules *d*_*k*_ start to approximate *d*^*\**^, then we may expect that the value of e.g., 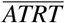 should converge to 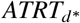. Previous theoretical work in this area suggests that we might expect small bias in large samples, but incorrect standard error estimation ^20,19^. We evaluate this claim via simulation in Section 4 below.

## 3 ESTIMATION

### 3.1 Learning treatment rules

The first task in estimation is to utilize data from each training sample to develop a treatment rule. To do this, we follow the Doubly-Robust (DR)-Learner approach outlined in Kennedy (2023) ^12^. This approach itself relies on cross-validation and thus our analysis considers *nested* cross-validation, wherein the *k*-th training sample is partitioned into *V* nested folds. The *v*-th nested training fold is used to fit certain *nuisance regressions* including a regression of (i) the outcome *Y* on antibiotic use *A* and covariates *W*; (ii) the indicator of antibiotic use *A* on covariates *W*; and (iii) the indicator of having a missing outcome Δ on *A* and *W*. We denote the outcome regression by *µ*(*a, w*) = *E*[*Y* | Δ = 1, *A* = *a, W* = *w*], the antibiotic regression by *π*(*a* | *w*) = *P*(*A* = *a* | *W* = *w*), and the missingness regression by *γ*(1 | *a, w*) = *P*(Δ = 1 | *A* = *a, W* = *w*) and denote estimates of these quantities developed using the *v*-th training sample nested in the *k*-th training sample by 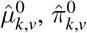, and 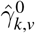, respectively.

In general any regression approach could be used to estimate these regression. In our analysis, we relied on Super Learner, an ensemble machine learning approach, to estimate each of these regressions ^13^.

Once the nuisance regression estimates are obtained, for each observation *i* in the *v*-th validation sample nested in the *k*-th training sample, we define the pseudo-outcome

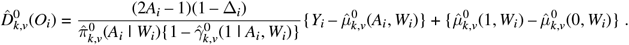

Next, using data from the *v*-th validation sample nested in the *k*-th training sample, we fit a regression of the pseudo-outcome 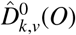 on *Z*. We denote this fitted regression by 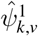. This approach is referred to as doubly robust, since it has been shown that that if either (i) the outcome regression or (ii) the antibiotic regression and the missingness regression are consistent for their respective outcomes, then the conditional mean of 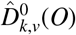 given *Z* converges in an appropriate sense to the CATE, *ψ*(*Z*) ^12^.

We repeat this process of using the *v*-th training sample to estimate nuisance parameters and the *v*-th validation to evaluate regression of the pseudo-outcome onto *Z* for each of the *V* nested cross-validation folds, resulting in CATE estimates 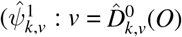. We average these estimates to define a treatment rule based on the *k*-th training sample by letting 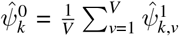 and 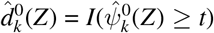.

### 3.2 Doubly Robust Estimators for Effect Estimates

Given treatment rule 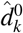 from the *k*-th training sample, we derive our estimates of parameters evaluating the rule as follows. First, we let 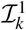 denote the set of indices of observations in the *k*-th validation sample and let 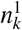 denote the number of observations in the *k*-th validation sample. The estimate *p*_*k*_ is given by 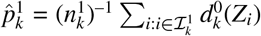, the sample proportion of observations in the *k*-th validation sample that would be treated under rule 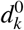. To estimate the effect parameters of interest, we utilize cross-fitted one-step estimators ^21^. To compute these estimators, we first re-estimate the nuisance regressions using the entire *k*-th training sample (as opposed to the nested training samples used to derive treatment rules above) and let 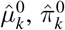, and 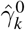 denote these estimates. Our estimate of *θ*_*k*_(*a* | *d*) for a particular (*a, d*) *∈* {0, 1}^2^, is constructed by defining for each 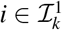

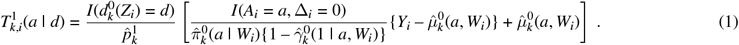

The final estimate of *θ*_*k*_(*a* | *d*) is

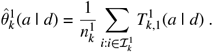

Under standard assumptions 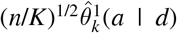 will have an asymptotically Normal distribution. Briefly, these assumptions include that the relevant nuisance regression estimates 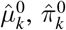 and 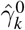 converge to their respective targets with respect to an appropriate norm at an appropriately fast rate. Because treatment assignment in the ABCD is randomized, the estimate 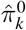 should satisfy these rate conditions by design. However, we still require sufficiently fast rates of estimation for the outcome model and missingness models. This motivates our usage of super learner. By including a diverse set of learners as candidates in the super learning procedure, we hope to uncover at least one regression algorithm that achieves an appropriately fast rate of convergence. The second assumption needed for asymptotic Normality is that the random variable 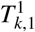 converges in an appropriate norm to a random variable *T*_*k*_, which is defined by replacing estimates of nuisance regressions with their respective true quantities in (1). Notably, because we are constructing our effect estimates using nuisance regressions learned in the training samples, we do not require so-called Donsker conditions to establish asymptotic Normality (see e.g., ^22^). This is akin to sample-splitting used by so-called “double machine learning estimators” of causal effects ^23^.

The asymptotic variance of 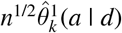 can be estimated using

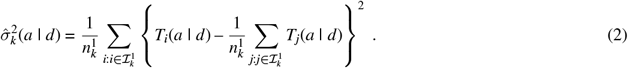

Finally, to obtain an overall estimate, we average the *k*-specific estimates over the *K* cross-validation folds. Hubbard et al. ^19^ provide assumptions under which the resulting estimator will have an approximately Normal sampling distribution. A flowchart depicting the overall estimation procedure can be seen in Figure 1.

**FIGURE 1.**
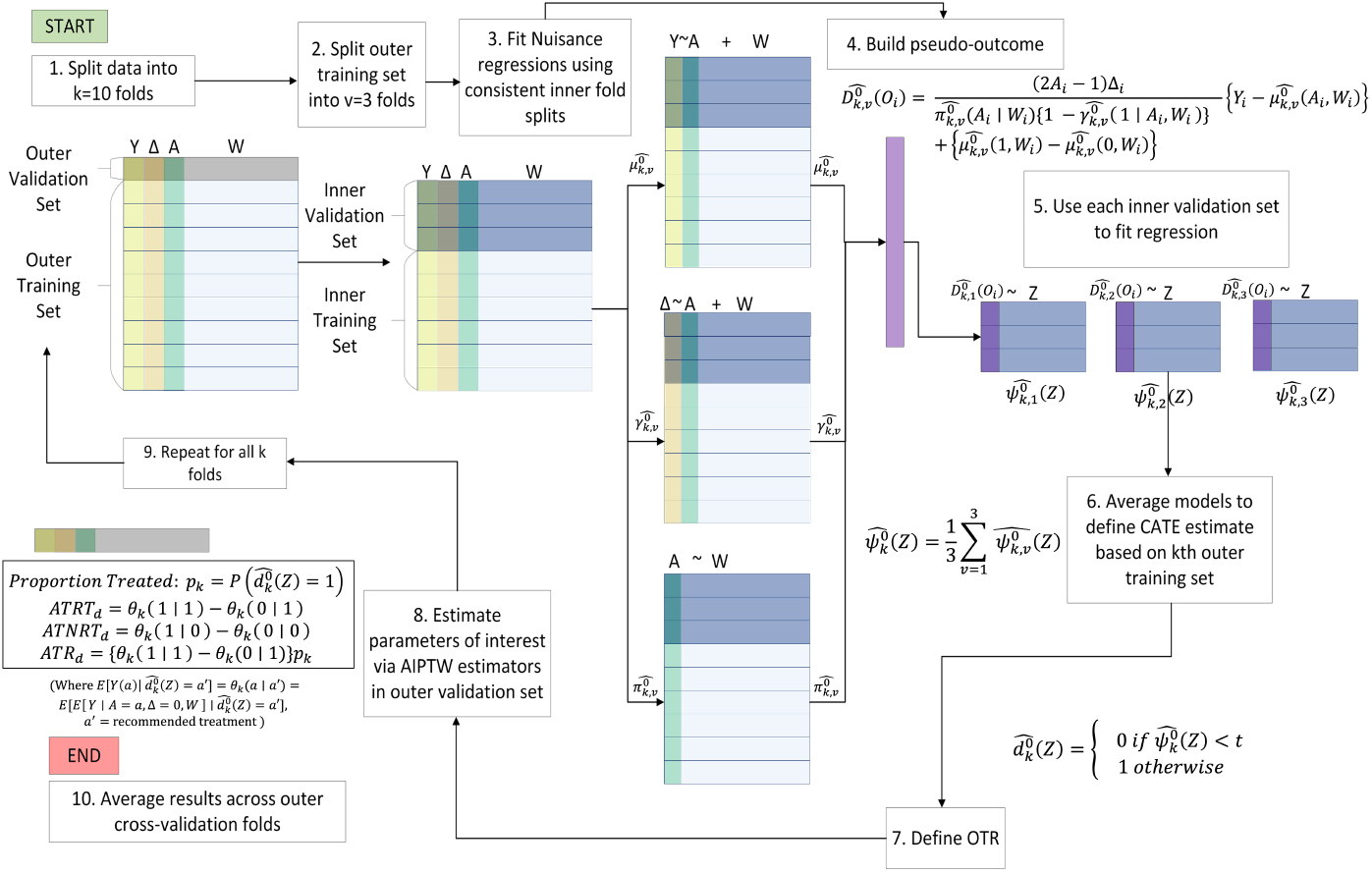
Estimation procedure for doubly-robust treatment rule with nested cross validation

A benefit of using doubly robust estimators is that inference for effects is facilitated through straightforward application of the delta method for influence functions ^24^. Specifically, for each observation in the *k*-th validation sample, we define the vector 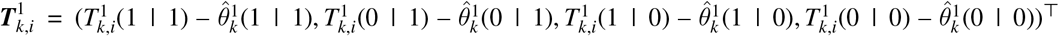. We define 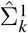 to be the sample covariance of 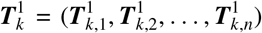. With this covariance matrix in hand, simple delta method calculus yields standard error estimates for each of the effects of interest. For example, let ***A*** = (1, –1, 0, 0)^*⊤*^. The standard error estimate of the estimate of 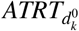 is 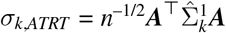 and an asymptotic Wald-style 95% confidence interval is given by 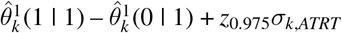, where *z*_0.975_ is the 0.975-quantile of a standard Normal distribution.

Similar delta method calculus can be used to derive the standard errors for the comparison of these effects between different rules. For example, suppose we are interested in comparing the ATRT between rules 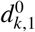 and 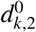 developed in the *k*-th training sample based on input covariates *Z*_1_ and *Z*_2_, respectively. We first define a rule-specific vector 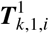 for rule 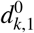 and 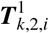 for rule 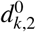. To obtain the covariance matrix we simply stack these vectors together; allowing for a slight abuse of notation, we define this stacked vector as 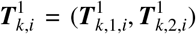 and define 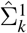 to be the eight-by-eight sample covariance of the stacked vector. Again, with access to this covariance matrix estimate, straightforward delta method calculus yields standard errors and confidence intervals for a comparison of the effects of interest. For example, to compare the ATRT between rules 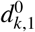 and 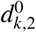, we may define ***B*** = (1, –1, 0, 0, –1, 1, 0, 0)^*⊤*^ and obtain a standard error estimate for the comparison as 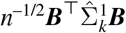, which can be used to create a Wald-style confidence interval as above.

## 4 SIMULATION STUDY

### 4.1 Data Generation

We designed a simulation study to evaluate our method based on the observed ABCD trial data. A list of all covariates, outcomes, and treatment recorded in the dataset can be found in table 1. We simulated our data such that each variable has a marginal distribution that closely follows the observed data. The majority of covariates were simulated independently for simplicity, while malnutrition-related covariates were simulated to have dependencies based on epidemiologically plausible relationships. Details on the distributions of each simulated covariate can be found in supplement sections 1.1-1.2. Assignment to azithromycin was drawn from a Bernoulli distribution with success probability 0.5.

We chose to focus on length-for-age z-score at day 90 of the trial as our primary outcome measure in the simulation study. We ensured the outcome model in our simulation was reflective of the real data by using a model fit the the real dataset to inform the covariates we include and their associated coefficients. We began by using LASSO with ten-fold cross-validated selection of the shrinkage parameter in the placebo arm of the real dataset. The LASSO model selected duration of diarrhea prior to enrollment, length for age z-score at enrollment, study site, and socioeconomic status quintile as variables to include in the model. Next, we added additional biologically plausible terms to the model including age and a binary indicator for *Shigella* presence. We further incorporated interaction terms with the binary treatment covariate *A*_*azithromycin*_. These interaction terms include (1) an interaction between *Shigella* and azithromycin; (2) an interaction between *Shigella*, length-for-age z-score at baseline, and azithromycin; and (3) an interaction between *Shigella*, length-for-age z-score, age, and azithromycin. The coefficients of these cross-product terms were selected such that (i) there was a non-zero treatment effect of azithromycin only if Shigella was present; (ii) the treatment effect was stronger for children with lower baseline length-for-age *Z* score; (iii) the magnitude of effect modification of treatment by baseline length-for-age *Z* score was lessened for older children. This reflects the biologically plausible scenario wherein the children who benefit most from antibiotic treatment are those who are youngest and with the worst nutrition, while malnutrition is a weaker determinant of treatment benefit in older-aged children. The resulting model used to simulate *Y*_*lazd*90_ is detailed in supplemental equation 1.3. Outcomes are missing completely at random for 4% of children.

### 4.2 Treatment rules of interest

Using the data generation process described above, we evaluated our proposed estimates of our effect estimands of interest under treatment rules based on two different sets of covariates: (1) a “comprehensive” set of covariates (with associated rule denoted by *d*_*c*_), which included all measured diagnostic, child, and clinical characteristics, and (2) a “host-only” set of covariates (with associated rule denoted by rule *d*_*h*_) that included only covariates related to growth and sociodemographic characteristics of the child. We also evaluated rules developed using several different thresholds. To select the thresholds, we computed the population distribution of conditional average treatment effects (Figure 2). Recall that data were generated such that only children with *Shigella* had a benefit of treatment. As a result, the CATE based on the “host-only” covariates tended to be smaller as subgroups defined by host-only covariates included both children with and without *Shigella*. Thus, we evaluated treatment rules based on the comprehensive set of covariates that thresholded CATE estimates at *t* = 0.05, 0.15, 0.25, 0.35 z-scores, while we evaluated host-only rules that thresholded CATE estimates at *t* = 0.025, 0.05, 0.075, 0.10 z-scores. Each range of thresholds was selected to capture scenarios where as few as 0.1% to as many as 99% of children are expected to be treated under each rule, thereby providing a range of effective sample sizes for evaluating our estimators.

**FIGURE 2.**
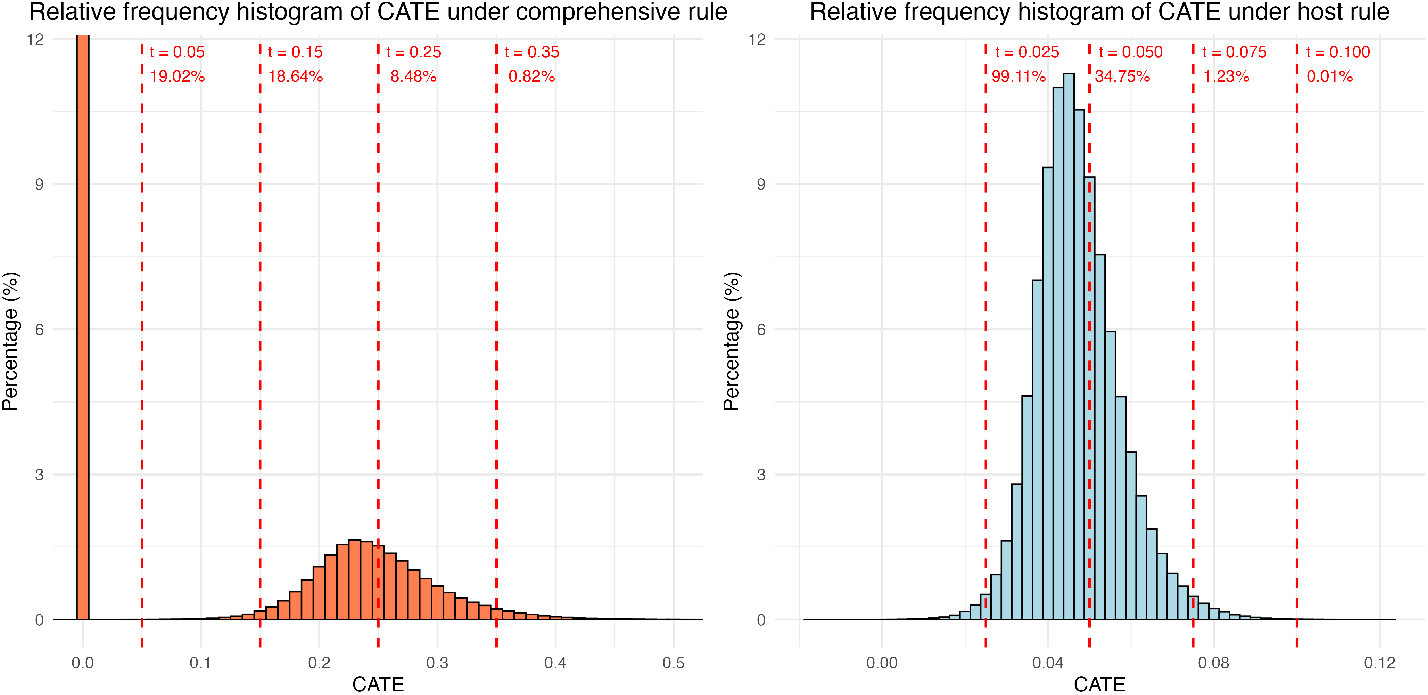
Population distribution of conditional average treatment effects under comprehensive and host rules with chosen thresholds and true proportions treated

For each rule (defined by choice of covariates and threshold), we estimated the effect parameters of interest. We also estimated the difference in estimated effects under the rules based on different covariates at their common threshold of *t* = 0.05. For each data set the true value of the data-adaptive effect parameters were calculated by simulating a large independent test data set and using these data to numerically approximate the true value of the parameters. We used these values to compute bias and assessed 95% confidence interval coverage for our estimates. In addition, we used the test data set to compute expected effects under a treatment rule based on the true, rather than estimated, CATE for each set of covariates and each threshold. The effects under these oracle rules can be found in table 2.

**TABLE 2.** True values of effect parameters under oracle treatment rules developed based on the CATE given a comprehensive set of input covariates and given host-only input covariates

| Covariate set | Parameter | $t = 0.05$ | $t = 0.15$ | $t = 0.25$ | $t = 0.35$ |
| --- | --- | --- | --- | --- | --- |
| Comprehensive | $P(d_c^*(Z) = 1)$ | 0.1901 | 0.1863 | 0.0845 | 0.0082 |
| | $ATRT_{d_c}$ | 0.2481 | 0.2498 | 0.2948 | 0.3776 |
| | $ATNRT_{d_c}$ | -0.0026 | -0.0018 | 0.0224 | 0.0424 |
| | $ATR_{d_c}$ | 0.0472 | 0.0465 | 0.0249 | 0.0031 |
| | $ATRT_{d_c} - ATNRT_{d_c}$ | 0.2507 | 0.2516 | 0.2724 | 0.3352 |
| Covariate set | Parameter | $t = 0.025$ | $t = 0.050$ | $t = 0.075$ | $t = 0.100$ |
| Host-only | $P(d_h^*(Z) = 1)$ | 0.9911 | 0.3475 | 0.0123 | 0.0001 |
| | $ATRT_{d_h}$ | 0.0452 | 0.0549 | 0.0774 | 0.1465 |
| | $ATNRT_{d_h}$ | 0.0031 | 0.0408 | 0.0447 | 0.0450 |
| | $ATR_{d_h}$ | 0.0045 | 0.0019 | 0.0000 | 0.0000 |
| | $ATRT_{d_h} - ATNRT_{d_h}$ | 0.0421 | 0.0141 | 0.0327 | 0.1015 |

**TABLE 3.** Simulation results for comprehensive and host rules (n = 6692, replicates = 1000)

| Rule | Effect Estimate | Performance Metric | Data Adaptive |  | Oracle |  |
| --- | --- | --- | --- | --- | --- | --- |
| | | | $t = 0.05$ | $t = 0.25$ | $t = 0.05$ | $t = 0.25$ |
| **Comprehensive** | $P(d_c^*(Z) = 1)$ | Bias | -0.001 | 0.000 | 0.171 | -0.053 |
|  |  | 95% CI Coverage | 0.924 | 0.935 | 0.001 | 0.012 |
| | $ATRT_{d_c}$ | Bias | -0.006 | -0.001 | -0.109 | -0.058 |
|  |  | 95% CI Coverage | 0.927 | 0.930 | 0.084 | 0.940 |
| | $ATNRT_{d_c}$ | Bias | -0.004 | -0.005 | 0.004 | 0.018 |
|  |  | 95% CI Coverage | 0.929 | 0.925 | 0.944 | 0.801 |
| | $ATR_{d_c}$ | Bias | -0.002 | 0.000 | -0.001 | -0.017 |
|  |  | 95% CI Coverage | 0.932 | 0.961 | 0.912 | 0.062 |
| | $ATRT_{d_c} - ATNRT_{d_c}$ | Bias | -0.002 | 0.004 | -0.113 | -0.076 |
|  |  | 95% CI Coverage | 0.935 | 0.922 | 0.156 | 0.924 |
| | | | $t = 0.05$ | $t = 0.075$ | $t = 0.05$ | $t = 0.075$ |
| **Host** | $P(d_h^*(Z) = 1)$ | Bias | 0.000 | 0.000 | 0.125 | 0.245 |
|  |  | 95% CI Coverage | 0.936 | 0.943 | 0.046 | 0.000 |
| | $ATRT_{d_h}$ | Bias | -0.005 | -0.005 | -0.007 | -0.028 |
|  |  | 95% CI Coverage | 0.931 | 0.915 | 0.936 | 0.898 |
| | $ATNRT_{d_h}$ | Bias | -0.005 | -0.005 | 0.005 | 0.001 |
|  |  | 95% CI Coverage | 0.910 | 0.930 | 0.947 | 0.956 |
| | $ATR_{d_h}$ | Bias | -0.002 | -0.001 | 0.003 | 0.012 |
|  |  | 95% CI Coverage | 0.918 | 0.902 | 0.774 | 0.651 |
| | $ATRT_{d_h} - ATNRT_{d_h}$ | Bias | 0.000 | 0.000 | -0.013 | -0.030 |
|  |  | 95% CI Coverage | 0.896 | 0.911 | 0.903 | 0.871 |

**TABLE 4.** Simulation results for comprehensive vs host rule comparison for *t* = 0.05

| Effect Estimate | Performance Metric | Data Adaptive | Oracle |
| --- | --- | --- | --- |
| $ATRT_{d_c} - ATRT_{d_h}$ | Bias | -0.001 | -0.170 |
|  | 95% CI Coverage | 0.930 | 0.070 |
| $ATNRT_{d_c} - ATNRT_{d_h}$ | Bias | 0.002 | 0.067 |
|  | 95% CI Coverage | 0.915 | 0.309 |
| $ATR_{d_c} - ATR_{d_h}$ | Bias | 0.000 | -0.005 |
|  | 95% CI Coverage | 0.915 | 0.441 |

### 4.3 Procedure

We simulated 1000 datasets of size *n* = 6692 (the size of the true ABCD dataset) and applied our method to each one to obtain estimates and 95% confidence intervals for each effect of interest. We calculated bias and coverage of the nominal 95% confidence intervals for the both effects under the data-adaptive rules, as well as for oracle rules defined based on the true unknown CATE.

We included a range of candidate learners to estimate each nuisance parameter and for the doubly robust estimator of the CATE. These included default regression algorithms included in the SuperLearner R package ^25^, as well as custom learners created specifically based on biologically plausible hypotheses. The outcome learners included generalized linear model, random forest, multivariate adaptive regression splines, lasso and elastic-net regularized generalized linear models, gradient boosting, and eleven biologically plausible generalized linear models. The set of learners used to estimate the treatment probability was simpler given the randomized design and included only the sample mean and a generalized linear model. The set of learners used to estimate the probability of missingness was also simpler and included mean and two logistic regression models that explicitly modeled study site and calendar time to account for known patterns of missingness based on when outcome measurements were made during the course of the study. The CATE learners for both comprehensive and host rules included multivariate adaptive regression splines, random forest, lasso and elastic-net regularized generalized linear models, and a main-terms linear model. Additional details on the candidate Super Learner libraries can be found in Supplementary Table 5.

### 4.4 Results

Below, we present results for a subset of thresholds, with results for the remaining thresholds available in supplementary tables 6-7. Overall, we found that the proposed method exhibited low bias and approximately 95% coverage for each data-adaptive effect estimand of interest. This was observed across all thresholds and under both the host and comprehensive rules. Bias remained consistently low and coverage was near the nominal level across all thresholds, indicating robust performance even with small effective sample sizes. As expected, using our estimates for the purpose of deriving inference about effects under the oracle treatment rules created based on the unknown true CATE for a given threshold tended to yield biased inference with coverage often far below nominal levels.

**TABLE 5.** Data analysis for length-for-age z-score outcome aggregated across 5 seeds for host and comprehensive rules

| Effect Estimate | Threshold | Host Rule |  | Comprehensive Rule |  |
| --- | --- | --- | --- | --- | --- |
|  |  | Point Estimate | 95% CI | Point Estimate | 95% CI |
| $P(d^*(Z) = 1)$ | 0.06 | 0.40 | [0.39, 0.41] | 0.39 | [0.38, 0.40] |
|  | 0.08 | 0.26 | [0.25, 0.27] | 0.21 | [0.20, 0.22] |
|  | 0.10 | 0.18 | [0.17, 0.19] | 0.11 | [0.10, 0.12] |
| $ATRT_d$ | 0.06 | 0.05 | [-0.01, 0.11] | 0.05 | [0.01, 0.10] |
|  | 0.08 | 0.02 | [-0.15, 0.20] | 0.05 | [-0.01, 0.12] |
|  | 0.10 | -0.02 | [-0.27, 0.23] | 0.06 | [-0.04, 0.17] |
| $ATNRT_d$ | 0.06 | 0.06 | [0.02, 0.09] | 0.05 | [0.02, 0.09] |
|  | 0.08 | 0.05 | [0.02, 0.09] | 0.05 | [0.02, 0.08] |
|  | 0.10 | 0.06 | [0.03, 0.09] | 0.05 | [0.02, 0.08] |
| $ATR_d$ | 0.06 | 0.02 | [0.00, 0.03] | 0.02 | [0.00, 0.04] |
|  | 0.08 | 0.01 | [0.00, 0.03] | 0.01 | [0.00, 0.02] |
|  | 0.10 | 0.01 | [-0.01, 0.02] | 0.01 | [0.00, 0.02] |
| $ATRT_d - ATNRT_d$ | 0.06 | 0.00 | [-0.07, 0.07] | 0.00 | [-0.05, 0.06] |
|  | 0.08 | -0.03 | [-0.21, 0.15] | 0.00 | [-0.07, 0.07] |
|  | 0.10 | -0.08 | [-0.33, 0.17] | 0.01 | [-0.10, 0.13] |

**TABLE 6.** Comparison of comprehensive - host rules for length-for-age z-score at day ninety

| Effect Estimate | Threshold | Value | 95% Confidence Interval |  |
| --- | --- | --- | --- | --- |
|  |  |  | Lower Bound | Upper Bound |
| $ATRT_{dc} - ATRT_{dh}$ | 0.06 | -0.006 | -0.050 | 0.038 |
|  | 0.08 | -0.012 | -0.083 | 0.059 |
|  | 0.10 | 0.005 | -0.143 | 0.152 |
| $ATNRT_{dc} - ATNRT_{dh}$ | 0.06 | 0.005 | -0.022 | 0.033 |
|  | 0.08 | 0.000 | -0.019 | 0.018 |
|  | 0.10 | -0.009 | -0.022 | 0.004 |
| $ATR_{dc} - ATR_{dh}$ | 0.06 | -0.006 | -0.023 | 0.010 |
|  | 0.08 | -0.003 | -0.017 | 0.011 |
|  | 0.10 | 0.005 | -0.007 | 0.017 |

**TABLE 7.** Data analysis for day three diarrhea outcome aggregated across 5 seeds for host and comprehensive rules

| Effect Estimate | Threshold | Host Rule |  | Comprehensive Rule |  |
| --- | --- | --- | --- | --- | --- |
|  |  | Point Estimate | 95% CI | Point Estimate | 95% CI |
| $P(d^*(Z) = 1)$ | -0.06 | 0.39 | [0.38, 0.40] | 0.39 | [0.38, 0.40] |
|  | -0.08 | 0.20 | [0.19, 0.21] | 0.22 | [0.21, 0.23] |
|  | -0.10 | 0.10 | [0.10, 0.11] | 0.12 | [0.11, 0.13] |
| $ATRT_d$ | -0.06 | -0.05 | [-0.09, -0.01] | -0.09 | [-0.13, -0.05] |
|  | -0.08 | -0.06 | [-0.16, 0.03] | -0.10 | [-0.15, -0.05] |
|  | -0.10 | -0.03 | [-0.21, 0.16] | -0.10 | [-0.18, -0.03] |
| $ATNRT_d$ | -0.06 | -0.06 | [-0.09, -0.03] | -0.03 | [-0.06, 0.00] |
|  | -0.08 | -0.05 | [-0.08, -0.03] | -0.04 | [-0.06, -0.01] |
|  | -0.10 | -0.05 | [-0.08, -0.03] | -0.05 | [-0.07, -0.02] |
| $ATR_d$ | -0.06 | -0.02 | [-0.03, 0.00] | -0.03 | [-0.05, -0.02] |
|  | -0.08 | -0.01 | [-0.02, 0.00] | -0.02 | [-0.03, -0.01] |
|  | -0.10 | 0.00 | [-0.01, 0.00] | -0.01 | [-0.02, 0.00] |
| $ATRT_d - ATNRT_d$ | -0.06 | 0.01 | [-0.04, 0.06] | -0.06 | [-0.11, -0.01] |
|  | -0.08 | -0.01 | [-0.11, 0.09] | -0.06 | [-0.12, 0.00] |
|  | -0.10 | 0.03 | [-0.16, 0.21] | -0.06 | [-0.13, 0.02] |

**TABLE 8.**
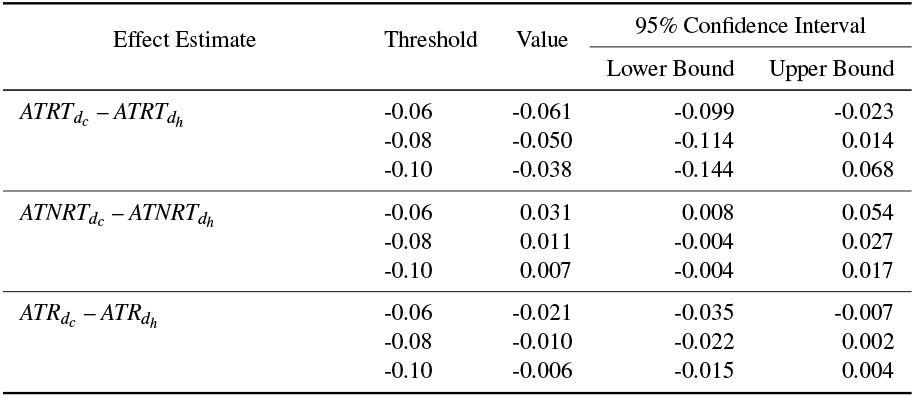
Comparison of comprehensive - host rules for diarrhea at day three

Our method also demonstrates low bias and nominal coverage when comparing the data-adaptive effect estimands between the two rules developed based on different sets of input covariates. As with the effect estimands for a given set of covariates, inference regarding the oracle parameters yielded biased results with poor coverage.

## 5 DATA ANALYSIS

We studied the same comprehensive and host rules evaluated in the simulation dataset using the real ABCD dataset for the length-for-age z-score at day ninety outcome. We also examined comprehensive and host rules for the binary outcome of diarrhea at day three post-enrollment. The same set of Super Learner libraries were used for the nuisance regressions, while an expanded set of libraries was used for the CATE models (see supplementary table 5). Rules were derived by thresholding CATE estimates at *t* = 0.06, 0.08, and 0.10 z-scores for the length-for-age z-score outcome, and *t* = –0.06, –0.08, –0.10 risk difference for the day three diarrhea outcome. The entire estimation process was repeated five times and results were aggregated across runs to account for variability in the cross-validation and machine learning procedures ^26^.

### 5.1 Length-for-age z-score at day ninety

#### 5.1.1 Rules based on host-only covariates

At a threshold of 0.06, the treatment rules based on host characteristics treat 39.8% (95% CI: 38.6% to 40.9%) of the population on average. The average proportion recommended treatment decreased to 26.3% (95% CI: 25.2% to 27.3%) using a threshold of 0.08 and 17.6% (95% CI: 16.7% to 18.6%) using a threshold of 0.10. Focusing on the threshold 0.06, azithromycin is expected to improve the length-for-age z-score by 0.05 (95% CI: -0.01 to 0.11) compared to placebo among children recommended for treatment by the derived treatment rules. This is about the same as the expected improvement for children *not* recommended treatment under the derived rules such that 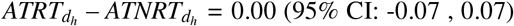, suggesting that a rule based on host characteristics at a threshold of 0.06 z-score does not effectively distinguish between children in terms of their expected benefit. Implementation of the rules at a population level is expected to improve length-for-age z-score by 0.02 (95% CI: 0.00 to 0.03) on average.

#### 5.1.2 Rules based on a comprehensive set of covariates

The treatment rules derived based on a comprehensive collection of covariates treat 39.2% (95% CI: 38.0% to 40.3%) of the population on average at a threshold of 0.06. Like the host rule, proportion treated decreases as the threshold increases with 21.2% (95% CI: 20.2% to 22.2%) at a threshold of 0.08 and 10.9% (95% CI: 10.1% to 11.6%) at a threshold of 0.10. There was no evidence of a difference between 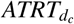 and 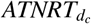, with their differences estimated to be 0.00 (95% CI: -0.05 to 0.06). This indicates that expanding the set of information available to the treatment rule to not discernibly impact our ability to distinguish between children who would and would not benefit from treatment. Population-level effect is the same as the host-rule with 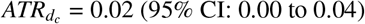.

#### 5.1.3 Comparing rules

We found that rules derived based on host-only characteristics tended to have nearly identical performance as those based on a more comprehensive set of covariates across all thresholds. The difference in estimates between comprehensive and host rules were close to zero, providing no evidence that there is benefit to incorporating more comprehensive information into treatment rules, as it relates to identifying children with larger expected benefits in terms of growth. This, paired with the lack of heterogeneity in average treatment benefit between the *ATRT*_*d*_ and *ATNRT*_*d*_, suggests benefit of azithromycin for linear growth is relatively uniform across the population. While there is evidence to support benefit of azithromycin treatment on linear growth at a population level, there is no clear evidence that a subgroup of children who would benefit more from this treatment can be identified.

### 5.2 Diarrhea at day three post-enrollment

#### 5.2.1 Rules based on host-only covariates

At a threshold of 6% risk reduction for day three diarrhea, 38.7% (95% CI: 37.5% to 39.8%) of the population was treated on average. Like the host rule for the length-for-age z-score, the proportion of the population recommended treatment decreased as the treatment threshold increased in magnitude. Treatment in accordance to the rule decreased the risk of diarrhea three days after enrollment by 4.7% (95% CI: 1.0% to 8.5%) on average among children recommended treatment by the derived rules. As with the host characteristic rule for length-for-age z-score, there was not a significant difference between children recommended and not recommended treatment under the rule with regards to their expected benefit 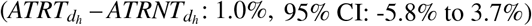 to 3.7%).

#### 5.2.2 Rules based on comprehensive set of covariates

Similarly to the host rule, 39.2% (95% CI: 38.0% to 40.3%) of the population was treated on average under the comprehensive rule at a threshold of 6% risk reduction. Risk of diarrhea three days after enrollment was decreased by 9.1% (95% CI: 12.9% to 5.3%) on average in the subgroup that was recommended treatment. Unlike the host rule, the comprehensive rule effectively distinguished between children in terms of their expected benefit with an average difference in 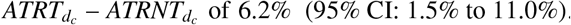. At a population level, implementation of a comprehensive rule reduced risk of diarrhea at day three by 3.4% (95% CI: 1.9% to 4.8%).

At the 10% risk reduction threshold, only 10.5% (95% CI: 9.7% to 11.2%) of the population was treated on average compared to 39.2% at the 6% threshold. The average risk reduction in the subgroup recommended treatment under the rule was 10.3% (95% CI 3.2% to 17.5%), a larger risk reduction than in the subgroup recommended treatment at the 6% threshold. The average risk reduction at a population level of the comprehensive rule is smaller using this threshold at 1.0% (95% CI: 0.0% to 1.9%). This highlights the impact that adjusting the treatment threshold can have on both a subgroup and population level.

#### 5.2.3 Rule comparison

In contrast with our rules for length-for-age z-score outcome, the comprehensive rule had improved performance over a rule based solely on host characteristics for diarrhea at day three. For one set of simulated rules, the comprehensive rule had a 6.1% (95% CI: 2.3% to 9.9%) larger risk reduction than the rule based on host characteristics alone for the subgroup recommended treatment under each rule using a threshold of 6% risk difference. At the population level, the comprehensive rule decreased the risk of day three diarrhea by 2.1% (95% CI: 0.7% to 3.5%) more than the host rule at the same risk reduction threshold. This, taken with the observed heterogeneity in *ATRT*_*d*_ and *ATRNT*_*d*_, suggests azithromycin does have a larger benefit on diarrhea at day three in population subgroups. The comprehensive rule includes illness characteristics and pathogen diagnostics that are specific to the given diarrheal episode. Inclusion of these covariates in treatment rules for diarrhea at day three can help identify subsets of children who would benefit more from treatment than treatment rules based on host characteristics alone.

## 6 DISCUSSION

We applied a framework based on machine learning and doubly-robust estimation for the study of treatment rules for azithromycin in children with watery diarrhea. The doubly-robust properties of the DR-Learner and our effect estimates ensure accurate estimation of our effects of interest if the outcome model or the propensity and missingness models are correctly specified. The use of Super Learner ensemble modeling allows for an optimally weighted combination of candidate models, and the nested cross-validation procedure minimizes overfitting of these models. Our simulation results validated that the method is appropriate for inference on data-adaptive target parameters.

A unique feature of our work is the need to explicitly consider tradeoffs between the proportion of children treated under a rule and their expected magnitude of treatment benefit. We examined rules based on thresholding expected benefits, which allows policymakers to maintain a patient-focused approach to antibiotic treatment, while also balancing potential concerns about antibiotic resistance. Another strength of our approach is that we are able to formally compare the performance of different rules based on different sets of input covariates. Such comparisons are critical for identifying which data may be most important to prioritize in tailoring treatment recommendations for children. For example, our analysis showed evidence of improvement in the diarrhea duration outcome when additional characteristics beyond host factors were incorporated into the treatment rule. This suggests that collecting additional data such as point-of-care diagnostics could be useful for optimizing short-term outcomes.

The applicability of our work is somewhat limited by our reliance on data-adaptive target parameters. For drawing biological conclusions about mechanisms of treatments, it may be more appropriate to consider the performance of rules based on the unknown oracle treatment rule. On the other hand, the oracle treatment rule is never known in practice and therefore it may be entirely appropriate to consider only rules that are developed based on data, as these are the treatment rules that are actually feasible to implement in practice. Therefore, we suggest that inference on data-adaptive target parameters may give insight into how realistic machine learning-based treatment rules may perform if deployed in real world settings.

The need to compare the performance of rules based on different sets of covariate inputs has wide ranging applications outside the specific context of diarrheal illness in children. For example, treatment rules are often created based on specific biomarkers that may be expensive to measure or not widely available in all clinical settings. Our approach allows investigators to quantify the marginal value added by measuring these biomarkers in terms of the ability to discriminate between those who will and will not benefit from treatment. To meet the demand for these analyses, we have developed an R package that implements our analysis pipeline and allows for straightforward comparisons of such rules. The package is freely available on GitHub (https://github.com/allicodi/drotr).

## Data Availability

The de-identified data that support the findings of this study are available from the corresponding author [A.C.] upon reasonable request and signed data access agreement upon publication.

## ACKNOWLEDGMENTS

We thank the children who participated in this trial and their families as well as the physicians, pharmacists, and research assistants who helped conduct the trial in each of the 7 countries.

## ABCD STUDY GROUP MEMBERS

Dilruba Ahmed, Tahmeed Ahmed, Tahmina Alam, Hannah E. Atlas, Per Ashorn, Henry Badji, Mohamed Bakari, Naor Bar-Zeev, Rajiv Bahl, Aishwarya Chauhan, Fadima Cheick Haidara, Mohammod Jobayer Chisti, Jen Cornick, Flanon Coulibaly, Ayesha De Costa, Nigel Cunliffe, Saikat Deb, Emily L. Deichsel, Pratibha Dhingra, Usha Dhingra, Christopher P. Duggan, Queen Dube, Arup Dutta, Bridget Freyne, Wilson Gumbi, Jean Gratz, Eric R. Houpt, Aneeta Hotwani, Ohedul Islam, Vijay Kumar Jaiswal, Furqan Kabir, Mamun Kabir, Kevin Mwangi Kariuki, Irene Kasumba, Shaila S. Khan, Upendo Kibwana, Rodrick Kisenge, Karen L. Kotloff, Jie Liu, Victor Maiden, Dramane Malle, Karim Manji, Christine McGrath, Ashka Mehta, Cecylia Msemwa, Chifundo Ndamala, Latif Ndeketa, Churchil Nyabinda, Darwin Operario, Irin Parvin, Patricia B. Pavlinac, Jasnehta Permala-Booth, James A. Platts-Mills, Ira Praharaj, Farah Naz Qamar, Shahida Qureshi, Muhammad Waliur Rahman, Doreen Rwigi, Abraham Samma, Sunil Sazawal, Sadia Shakoor, Anil Kumar Sharma, Jonathon Simon, Benson O. Singa, Sarah Somji, Samba O. Sow, Christopher R. Sudfeld, Milagritos D. Tapia, Rozina Thobani, Caroline Tigoi, Stephanie N. Tornberg-Belanger, Aliou Toure, Judd L. Walson, Desiree Witte, and Mohammad Tahir Yousafzai.

## SUPPORTING INFORMATION

Additional supporting information may be found in the online version of the article at the publishers website.

## REFERENCES

1. Troeger C, Blacker BF, Khalil IA, et al. Estimates of the global, regional, and national morbidity, mortality, and aetiologies of diarrhoea in 195 countries: a systematic analysis for the Global Burden of Disease Study 2016. The Lancet Infectious Diseases. 2018;18(11):1211–1228. doi: 10.1016/S1473-3099(18)30362-1

2. Pavlinac PB, Platts-Mills JA, Liu J, et al. Azithromycin for Bacterial Watery Diarrhea: A Reanalysis of the AntiBiotics for Children With Severe Diarrhea (ABCD) Trial Incorporating Molecular Diagnostics. The Journal of Infectious Diseases. 2023:jiad252. doi: 10.1093/infdis/jiad252

3. Victora CG, Adair L, Fall C, et al. Maternal and child undernutrition: consequences for adult health and human capital. The Lancet. 2008;371(9609):340–357. Publisher: Elsevier doi: 10.1016/S0140-6736(07)61692-4

4. Richard SA, Black RE, Gilman RH, et al. Diarrhea in Early Childhood: Short-term Association With Weight and Long-term Association With Length. American Journal of Epidemiology. 2013;178(7):1129–1138. doi: 10.1093/aje/kwt094

5. Houpt ER, Ferdous T, Ara R, et al. Clinical Outcomes of Drug-resistant Shigellosis Treated With Azithromycin in Bangladesh. Clinical Infectious Diseases. 2021;72(10):1793–1798. doi: 10.1093/cid/ciaa363

6. Nie X, Wager S, others. Learning objectives for treatment effect estimation. arXiv preprint 1712.04912. 2017.

7. Künzel SR, Sekhon JS, Bickel PJ, Yu B. Metalearners for estimating heterogeneous treatment effects using machine learning. Proceedings of the national academy of sciences. 2019;116(10):4156–4165.

8. Kennedy EH. Towards optimal doubly robust estimation of heterogeneous causal effects. arXiv preprint 2004.14497. 2020.

9. Athey S, Imbens G. Recursive partitioning for heterogeneous causal effects. Proceedings of the National Academy of Sciences. 2016;113(27):7353–7360.

10. Wager S, Athey S. Estimation and inference of heterogeneous treatment effects using random forests. Journal of the American Statistical Association. 2018;113(523):1228–1242.

11. Nizam S, Codi A, Rogawski McQuade E, Benkeser D. Highly Adaptive Lasso for estimation of heterogeneous treatment effects and treatment recommendation. Journal of Causal Inference. 2024.

12. Kennedy EH. Towards optimal doubly robust estimation of heterogeneous causal effects. 2023. 2004.14497 [math, stat]

13. Laan v.dM, Polley E, Hubbard A. Super Learner. Statistical applications in genetics and molecular biology. 2007;6(1). doi: 10.2202/1544-6115.1309

14. Luedtke AR, Laan MJvd. Super-Learning of an Optimal Dynamic Treatment Rule. The international journal of biostatistics. 2016;12(1):305. Publisher: NIH Public Accessdoi: 10.1515/ijb-2015-0052

15. Montoya LM, Laan v.dMJ, Luedtke AR, Skeem JL, Coyle JR, Petersen ML. The optimal dynamic treatment rule superlearner: considerations, performance, and application to criminal justice interventions. The International Journal of Biostatistics. 2023;19(1):217–238. doi: 10.1515/ijb-2020-0127

16. Codi A. drotr: Doubly-robust methods for Optimal Treatment Rules. 2024. R package version 0.1.0.

17. The Antibiotics for Children With Diarrhea (ABCD) Study Group, Ahmed T, Chisti MJ, et al. Effect of 3 Days of Oral Azithromycin on Young Children With Acute Diarrhea in Low-Resource Settings: A Randomized Clinical Trial. JAMA Network Open. 2021;4(12):e2136726. doi: 10.1001/jamanetworkopen.2021.36726

18. Luedke AR, Laan v.dMJ. Optimal Individualized Treatments in Resource-Limited Settings. International Journal of Biostatistics. 2016.

19. Hubbard AE, Kherad-Pajouh S, Laan MJvd. Statistical Inference for Data Adaptive Target Parameters. The International Journal of Biostatistics. 2016;12(1):3–19. Publisher: De Gruyterdoi: 10.1515/ijb-2015-0013

20. Luedtke AR, Laan MJvd. Statistical inference for the mean outcome under a possibly non-unique optimal treatment strategy. The Annals of Statistics. 2016;44(2):713–742. Publisher: Institute of Mathematical Statisticsdoi: 10.1214/15-AOS1384

21. Pfanzagl J. Contributions to a General Asymptotic Statistical Theory. Lecture Notes in Statistics Springer New York, NY. 1 ed., 1982

22. Zheng W, Laan MJVD. Cross Validated Targeted Minimum-Loss-Based Estimation. In:,, Springer, 2011ch. 27:459–473

23. Chernozhukov V, Chetverikov D, Demirer M, et al. Double/debiased machine learning for treatment and structural parameters. The Econometrics Journal. 2018;21(1):C1–C68. doi: 10.1111/ectj.12097

24. Van Der Laan MJ, Rose S. Appendix A1: [Asymptotic Linearity: The Functional Delta Method]. In: Van Der Laan MJ, Rose S., eds. Targeted Learning: Causal Inference for Observational and Experimental Data,, Springer Series in Statistics. New York, NY: Springer, 2011:[521-524]

25. Polley E, LeDell E, Kennedy C, van der Laan M. SuperLearner: Super Learner Prediction. 2024. R package version 2.0-29.

26. Schader L, Song W, Kempker R, Benkeser D. Don’t Let Your Analysis Go to Seed: On the Impact of Random Seed on Machine Learning-based Causal Inference. Epidemiology (Cambridge, Mass.). 2024;35(6):764–778. doi: 10.1097/EDE.0000000000001782

