## Supplementary Material for "Machine learning for estimating and comparing clinical rules for treating diarrheal illness with antibiotics"

#### 1 Data Generation

##### 1.1 Dependently Simulated Covariates

$$
\begin{aligned}
\mu_{avemuac} &= 12.9818 - 0.3454 \times I(W_{SES} = \text{2nd quintile}) \\
&- 0.1088 \times I(W_{SES} = \text{3rd quintile}) + 0.1998 \times I(W_{SES} = \text{4th quintile}) \\
&- 0.3147 \times I(W_{SES} = \text{5th quintile}) + 0.3190 \times I(W_{sex} = \text{female}) \\
\mu_{lfazscore} &= -1.2366 - 0.4029 \times I(W_{SES} = \text{2nd quintile}) \\
&- 0.2715 \times I(W_{SES} = \text{3rd quintile}) + 0.3132 \times I(W_{SES} = \text{4th quintile}) \\
&- 0.4289 \times I(W_{SES} = \text{5th quintile}) - 0.2856 \times I(W_{sex} = \text{female}) \\
\mu_{wfazscore} &= -1.4689 - 0.5182 \times I(W_{SES} = \text{2nd quintile}) \\
&- 0.1286 \times I(W_{SES} = \text{3rd quintile}) + 0.2426 \times I(W_{SES} = \text{4th quintile}) \\
&- 0.4257 \times I(W_{SES} = \text{5th quintile}) - 0.1953 \times I(W_{sex} = \text{female}) \\
\mu_{wflzscore} &= -1.0507 - 0.3747 \times I(W_{SES} = \text{2nd quintile}) \\
&+ 0.0352 \times I(W_{SES} = \text{3rd quintile}) + 0.0943 \times I(W_{SES} = \text{4th quintile}) \\
&- 0.2382 \times I(W_{SES} = \text{5th quintile}) - 0.0772 \times I(W_{sex} = \text{female}) \\
\Sigma &= \begin{pmatrix} 1.3276 & 0.5344 & 1.0520 & 0.9529 \\ 0.5344 & 1.7310 & 1.1410 & 0.2196 \\ 1.0520 & 1.1410 & 1.4822 & 1.1427 \\ 0.9529 & 0.2196 & 1.1427 & 1.4796 \end{pmatrix}
\end{aligned}
$$

$$
W_{avemuac}, W_{lfazscore}, W_{wfazscore}, W_{wflzscore} \mid W_{sex}, W_{SES} \sim \text{MVN}\left(\mu = \begin{pmatrix} \mu_{avemuac} \\ \mu_{lfazscore} \\ \mu_{wfazscore} \\ \mu_{wflzscore} \end{pmatrix}, \Sigma = \Sigma\right) \tag{1}
$$

##### 1.2 Independently Simulated Covariates

Table 1: Table of independently simulated covariates $W$

| **Covariate (W)** | **Distribution** |
|---|---|
| Rotavirus Pathogen Quantity | $W_{Rotavirus} \sim Bin(n=1, p=0.24) \times \Gamma(shape=3.90, rate=1.25)$ |
| Norovirus Pathogen Quantity | $W_{Norovirus} \sim Bin(n=1, p=0.19) \times Unif(a=0, b=6)$ |
| Adenovirus Pathogen Quantity | $W_{Adenovirus} \sim Bin(n=1, p=0.19) \times \Gamma(shape=1.19, rate=0.43)$ |
| Astrovirus Pathogen Quantity | $W_{Astrovirus} \sim Bin(n=1, p=0.09) \times Unif(a=0, b=6)$ |
| Sapovirus Pathogen Quantity | $W_{Sapovirus} \sim Bin(n=1, p=0.17) \times \Gamma(shape=2.34, rate=0.93)$ |
| St. Etec Pathogen Quantity | $W_{ST\_ETEC} \sim Bin(n=1, p=0.23) \times Unif(a=0, b=7)$ |
| *Shigella* Pathogen Quantity | $W_{Shigella} \sim Bin(n=1, p=0.19) \times Weibull(shape=1.86, scale=3.06)$ |
| Campylobacter Pathogen Quantity | $W_{Campylobacter} \sim Bin(n=1, p=0.34) \times Weibull(shape=1.82, scale=2.73)$ |
| Tepec Pathogen Quantity | $W_{t\_EPEC} \sim Bin(n=1, p=0.22) \times Unif(a=0, b=6.5)$ |
| V. Cholerae Pathogen Quantity | $W_{V\_Cholerae} \sim Bin(n=1, p=0.03) \times \Gamma(shape=1.86, rate=1.06)$ |
| Salmonella Pathogen Quantity | $W_{Salmonella} \sim Bin(n=1, p=0.01) \times \Gamma(shape=1.28, rate=1.93)$ |
| Cryptosporidium Pathogen Quantity | $W_{Cryptosporidium} \sim Bin(n=1, p=0.20) \times \Gamma(shape=2.21, rate=0.77)$ |
| Child has vomited in current illness episode | $W_{vomit} \sim Bin(n=1, p=0.97)$ |
| Number of loose stools in 24-hour period prior to enrollment | $W_{lsstools} \sim Bin(n=7.06, p=0.51)$ |
| Number of solid stools in 24-hour period prior to enrollment | $W_{sstools} \sim Bin(n=0.08, p=0.43)$ |
| Number of days of diarrheal illness prior to enrollment | $W_{days\_diar} \sim Poisson(\lambda=2.5)$ |
| Level of dehydration | $P(W_{dehydration}=k) = \begin{cases} 0.45 & \text{if } k=1 \\ 0.5 & \text{if } k=2 \\ 0.05 & \text{if } k=3 \\ 0 & \text{otherwise} \end{cases}$ |
| Site | $W_{site} \sim DiscreteUnif(a=2, b=8)$ |
| Sex | $W_{sex} \sim Bin(n=1, p=0.5)$ |
| Age (in months) | $W_{age} \sim TruncatedGamma(range=(2,24), shape=4.88, rate=0.42)$ |
| Socioeconomic status quintile | $W_{SES} \sim DiscreteUnif(a=1, b=5)$ |
| Number of children < 5 years old in household | $W_{hh\_lt\_five} \sim TruncatedPoisson(\lambda=1.72, a=0)$ |
| Month enrolled | $W_{month} \sim DiscreteUnif(a=1, b=12)$ |

##### 1.3 Outcome Model

$$
\begin{aligned}
Y_{lazd90} = &-0.4213 + 0.0079 \times W_{dy1scrndiardays} + 0.1151 \times I(W_{site} = \text{Kenya}) \\
&- 0.0539 \times I(W_{site} = \text{Malawi}) + 0.0789 \times I(W_{site} = \text{Mali}) - 0.0250 \times I(W_{site} = \text{India}) \\
&- 0.1211 \times I(W_{site} = \text{Tanzania}) - 0.0059 \times I(W_{site} = \text{Pakistan}) \\
&- 0.0627 \times I(W_{SES} = \text{2nd quintile}) - 0.0246 \times I(W_{SES} = \text{3rd quintile}) \\
&+ 0.0214 \times I(W_{SES} = \text{4th quintile}) + 0.0518 \times I(W_{SES} = \text{5th quintile}) \\
&- 0.0002 \times W_{agemchild} + 0.8384 \times W_{lfazscore} - 0.0764 \times I(W_{Shigella} > 0) \\
&+ 0.1941 \times I(W_{Shigella} > 0) \times A_{azithromycin} \\
&- 0.0074 \times I(W_{Shigella} > 0) \times W_{lfazscore} \times A_{azithromycin} \\
&- 0.0025 \times I(W_{Shigella} > 0) \times W_{lfazscore} \times W_{agemchild} \times A_{azithromycin} + \epsilon
\end{aligned} \tag{2}
$$

$\epsilon \sim N(0, 0.5644)$

### 2 Candidate Super Learner Libraries

| **Outcome Model Libraries** |
|---|
| Generalized Linear Model (SL.glm) |
| Random Forest (SL.ranger) |
| Multivariate Adaptive Regression Splines (SL.earth) |
| Lasso and Elastic-Net Regularized Generalized Linear Models (SL.glmnet) |
| Gradient Boosting (SL.xgboost) |
| Generalized linear model with interaction for site and single pathogen quantity (rotavirus) |
| Generalized linear model with interaction for site and single pathogen quantity (*Shigella*) |
| Generalized linear model with interaction for site and rotavirus season |
| Generalized linear model with interactions for site and 2 pathogen quantities (rotavirus, *Shigella*) |
| Generalized linear model with interactions for site and pathogen quantities |
| Generalized linear model with interaction for site and vomiting |
| Generalized linear model with interactions for site, pathogen quantities, and illness characteristics |
| Generalized linear model with interactions for site, malnutrition indicators, and sociodemographic characteristics |
| Generalized linear model with interactions for site, illness characteristics, malnutrition indicators, and sociodemographic characteristics |
| Generalized linear model with interactions for site, pathogen quantities, illness characteristics, malnutrition indicators, and sociodemographic characteristics |

Table 2: Table of SuperLearner libraries used to fit outcome model. [1]

| **Treatment Model Libraries** |
|---|
| Mean (SL.mean) |
| Logistic regression model with all covariates |

Table 3: Table of SuperLearner libraries used to fit treatment model.

[1] Note: SL models are built into SuperLearner package. $W^1$ indicates all covariates in the dataset (see table **??** for full list). Covariates used in interaction terms are listed with name only for readability.

| **Missingness Model Libraries** |
|---|
| Mean (SL.mean) |
| Logistic regression with site, month |
| Logistic regression with site, month, and interaction |

Table 4: Table of SuperLearner libraries used to fit missingness model.

| **CATE Model Libraries** | | |
|---|---|---|
| **Study** | **Rule** | **Model** |
| **Simulation** | Comprehensive and host | Multivariate Adaptive Regression Splines (SL.earth) |
| | | Random Forest (SL.ranger) |
| | | Lasso and Elastic-Net Regularized Generalized Linear Models (SL.glmnet) |
| | | Generalized Linear Model (SL.glm) |
| **Real data** | Host | Multivariate Adaptive Regression Splines (SL.earth) |
| | | Random Forest (SL.ranger) |
| | | Lasso and Elastic-Net Regularized Generalized Linear Models (SL.glmnet) |
| | | Generalized Linear Model (SL.glm) |
| | | Gradient boosting (SL.xgboost) |
| | | Generalized linear model with terms for host characteristics (malnutrition indicators, sociodemographic characteristics) |
| | | Generalized linear model with terms for host characteristics with pairwise interactions |
| **Real data** | Comprehensive | Multivariate Adaptive Regression Splines (SL.earth) |
| | | Random Forest (SL.ranger) |
| | | Lasso and Elastic-Net Regularized Generalized Linear Models (SL.glmnet) |
| | | Generalized Linear Model (SL.glm) |
| | | Gradient boosting (SL.xgboost) |
| | | Generalized linear model with terms for host characteristics (malnutrition indicators, sociodemographic characteristics) |
| | | Generalized linear model with terms for host characteristics with pairwise interactions |
| | | Generalized linear model with terms for Shigella, $I(Shigella > 0)$ |
| | | Generalized linear model with terms for rotavirus, $I(rotavirus > 0)$ |
| | | Generalized linear model with all pathogen quantities and binary indicators for pathogen detection |
| | | Generalized linear model with all pathogen quantities and binary indicators for pathogen detection, pairwise interactions |
| | Generalized linear model with terms for illness characteristics | |
| | | Generalized linear model with terms for illness characteristics with pairwise interactions |
| | | Generalized linear model with terms for pathogens and illness characteristics |
| | | Generalized linear model with terms for pathogens and illness characteristics with pairwise interactions |
| | | Generalized linear model with all terms |
| | | Generalized linear model with all terms with pairwise interactions |

Table 5: Table of Super Learner libraries used to fit CATE models in simulation and real-data analyses

#### 3 Full simulation results

Table 6: Simulation results for comprehensive rule (n = 6692, replicates = 1000)

| | | t = 0.05 | | t = 0.15 | | t = 0.25 | | t = 0.35 | |
|---|---|---|---|---|---|---|---|---|---|
| Effect Estimate | Performance Metric | Oracle | Data Adaptive | Oracle | Data Adaptive | Oracle | Data Adaptive | Oracle | Data Adaptive |
| $P(d_c^*(Z) = 1)$ | Bias | 0.1708 | -0.0012 | -0.0443 | -0.0003 | -0.0534 | 0.0002 | -0.0024 | 0.0004 |
| | 95% CI Coverage | 0.0010 | 0.9240 | 0.1450 | 0.9340 | 0.0120 | 0.9350 | 0.3872 | 0.9564 |
| $ATRT_c$ | Bias | -0.1092 | -0.0059 | -0.0186 | -0.0035 | -0.0584 | -0.0006 | -0.1659 | -0.0218 |
| | 95% CI Coverage | 0.0840 | 0.9270 | 0.9250 | 0.9620 | 0.9400 | 0.9300 | 0.9231 | 0.9493 |
| $ATNRT_c$ | Bias | 0.0037 | -0.0038 | 0.0174 | -0.0045 | 0.0180 | -0.0048 | 0.0035 | -0.0046 |
| | 95% CI Coverage | 0.9440 | 0.9290 | 0.7620 | 0.9310 | 0.8010 | 0.9250 | 0.9464 | 0.9422 |
| $ATR_c$ | Bias | -0.0010 | -0.0021 | -0.0138 | -0.0008 | -0.0174 | 0.0002 | -0.0018 | 0.0001 |
| | 95% CI Coverage | 0.9120 | 0.9320 | 0.4150 | 0.9350 | 0.0620 | 0.9610 | 0.8140 | 0.9959 |
| $ATRT_c - ATNRT_c$ | Bias | -0.1129 | -0.0021 | -0.0360 | 0.0009 | -0.0764 | 0.0042 | -0.1694 | -0.0172 |
| | 95% CI Coverage | 0.1560 | 0.9350 | 0.8560 | 0.9590 | 0.9240 | 0.9220 | 0.9201 | 0.9504 |

Table 7: Simulation results for host rule (n = 6692, replicates = 1000)

| | | t = 0.025 | | t = 0.050 | | t = 0.075 | | t = 0.100 | |
|---|---|---|---|---|---|---|---|---|---|
| Effect Estimate | Performance Metric | Oracle | Data Adaptive | Oracle | Data Adaptive | Oracle | Data Adaptive | Oracle | Data Adaptive |
| $P(d_h^*(Z)=1)$ | Bias | -0.2982 | -0.0001 | 0.12500 | 0.00011 | 0.24532 | 0.00030 | 0.1310 | 0.0001 |
| | 95% CI Coverage | 0.0000 | 0.9340 | 0.04600 | 0.93600 | 0.00000 | 0.94300 | 0.0000 | 0.9440 |
| $ATRT_h$ | Bias | 0.0024 | -0.0051 | -0.00736 | -0.00537 | -0.02832 | -0.00493 | -0.0972 | -0.0072 |
| | 95% CI Coverage | 0.9250 | 0.9280 | 0.93600 | 0.93100 | 0.89800 | 0.91500 | 0.6780 | 0.9260 |
| $ATRNT_h$ | Bias | 0.0392 | -0.0099 | 0.00535 | -0.00543 | 0.00149 | -0.00541 | 0.0014 | -0.0054 |
| | 95% CI Coverage | 0.7970 | 0.9080 | 0.94700 | 0.91000 | 0.95600 | 0.93000 | 0.9500 | 0.9230 |
| $ATR_h$ | Bias | -0.0116 | -0.0033 | 0.00346 | -0.00242 | 0.01158 | -0.00113 | 0.0064 | -0.0006 |
| | 95% CI Coverage | 0.6830 | 0.9100 | 0.77400 | 0.91800 | 0.65100 | 0.90200 | 0.7380 | 0.9230 |
| $ATRT_h - ATRNT_h$ | Bias | -0.0368 | 0.0048 | -0.01271 | 0.00005 | -0.02981 | 0.00048 | -0.0986 | -0.0018 |
| | 95% CI Coverage | 0.7940 | 0.9010 | 0.90300 | 0.89600 | 0.87100 | 0.91100 | 0.6780 | 0.9200 |
